# Comorbidities associated with regional variations in COVID-19 mortality revealed by population-level analysis

**DOI:** 10.1101/2020.07.27.20158105

**Authors:** Hongxing Yang, Fei Zhong

## Abstract

Coronavirus disease 2019 (COVID-19), caused by severe acute respiratory syndrome coronavirus 2 (SARS-Cov-2), has developed into a global health crisis. Understanding the risk factors for poor outcomes of COVID-19 is thus important for successful management and control of the pandemic. However, the progress and severity of the epidemic across different regions show great differentiations. We hypothesized the origination of these differences are based on location-dependent variations in underlying population-wide health factors. Disease prevalence or incidence data of states and counties of the United States were collected for a group of chronic diseases, including hypertension, diabetes, obesity, stroke, coronary heart disease, heart failure, physical inactivation, and common cancers (e.g., lung, colorectal, stomach, kidney and renal). Correlation and regression analysis identified the prevalence of heart failure as a significant positive factor for region-level COVID-19 mortality. Similarly, the incidence of gastric cancer and thyroid cancer were also identified as significant factors contributing to regional variation in COVID-19 mortality. To explore the implications of these results, we re-analyzed the RNA-seq data for stomach adenocarcinoma (STAD) and colon carcinoma (COAD) from The Cancer Genome Atlas (TCGA) project. We found that expression of genes in the immune response pathways were more severely disturbed in STAD than in COAD, implicating higher probability for STAD patients or individuals with precancerous chronic stomach diseases to develop cytokine storm once infected with COVID-19. Taken together, we conclude that location variations in particular chronic diseases and cancers contribute significantly to the regional variations in COVID-19 mortality.

## Introduction

The pandemic caused by Coronavirus disease 2019 (COVID-19) is still ongoing. By the recent statistics of World Health Organization (WHO), this newly identified coronavirus has infected more than 15 million of people worldwide causing over six hundred thousand of deaths. It is estimated that about 15.7%-18.7% of COVID-19 patients could be classified as severe cases, requiring more intensive medical care and more likely to be fatal.^1–5^ Understanding the physiological, pathological, as well as demographic and socioeconomic factors associated with COVID-19 mortality is critical for decision makers to take effective and timely actions to reduce human life and economic losses and to alleviate the burden on healthcare workers.

Since the outbreak of the pandemic, great progress has been made in understanding the symptoms, risk factors, and prognosis of COVID-19. At present, important risk factors identified based on rapidly accumulating clinical observatory data include age (>65 years), sex (male), race, respiratory diseases, as well as other comorbidities, such as hypertension, obesity, diabetes, coronary heart diseases, acute myocardial infarction, heart failure, and malignance.^3,4,6–11^

The mortality of COVID-19 varies significantly across different countries and different administrative regions within a country (Figure 1). The variations could arise from various factors, including the strategies or interventions adopted by the administrations, demographic conditions, available public health resources, or the baseline health conditions of the populations. The prevalence/incidence of chronic diseases and cancers are important indicators for the baseline health conditions of a population and might have important influences on the progression or severity of a pandemic like COVID-19. To explore if the underlying health conditions could be correlated with or used as indicators for COVID-19 mortality, we performed a population analysis about the associations of chronic disease prevalence and cancer disease incidence with COVID19 mortality. Our analysis was focused on the states and counties of the United States of America (USA) as the epidemiological data is publicly available and well organized. Moreover, confining the analysis to regions within a single country also enabled control for unrelated socioeconomic or political factors such as law systems, or policy of central governments, although variations in local regulations or policies also exist among US states or counties.

**Figure 1.**
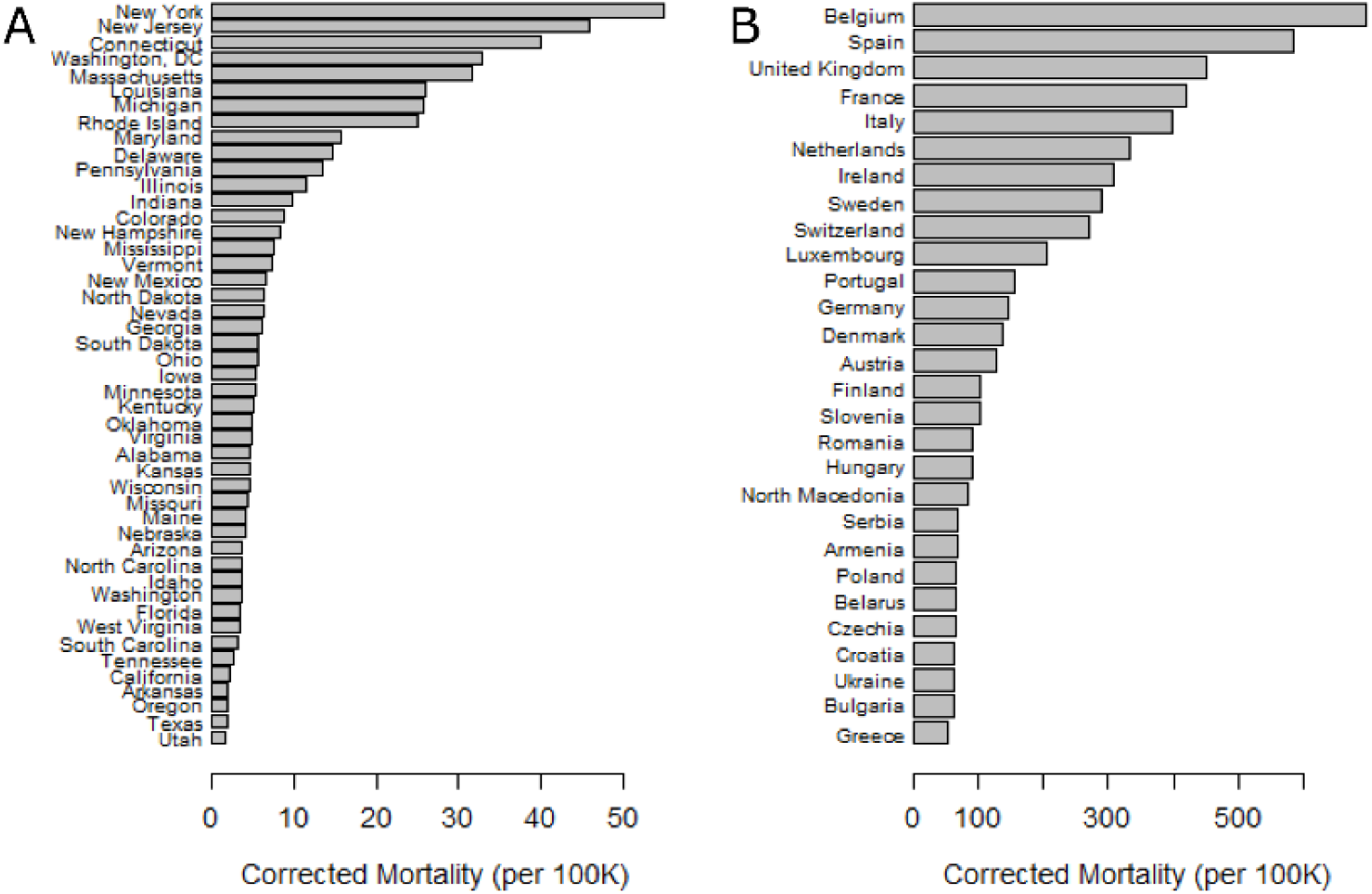
Variations of COVID-19 morality across states of USA (A) and European countries (B). Corrected mortality was the accumulated number of COVID-19-induced deaths on the 30^th^ day since the first day with at least 1 death in 100,000 people of a state or country.

## Data and Methods

### Data and analysis for prevalence or incidence of diseases

The prevalence data of chronic disease collected by The Behavioral Risk Factor Surveillance System (BRFSS) were downloaded from the Center for Disease Control and Prevent (CDC) webpage titled as ‘Division for Heart Disease and Stroke Prevention: Data Trends & Maps’, with the most recent year with available data being 2018. The cancer incidence data were retrieved from the CDC sub-website for ‘United States Cancer Statistics: Data and Visualizations’ (https://gis.cdc.gov/Cancer/USCS/DataViz.html), with the most recent year with available data being 2017. The COVID-19 mortality for counties of United States (US) and European countries were downloaded from the github site established by The Center for Systems Science and Engineering (CSSE) at John Hopkins University (https://github.com/CSSEGISandData). Daily death numbers for US states were obtained by aggregating daily death numbers of the counties belonging to each state. Corrected mortality for administrative regions was calculated as the accumulated number of COVID-19 induced deaths per 100,000 population on the 30^th^ day since the first day with at least 1 in 100,000 people of a region being observed.

Correlation and regression analysis were performed in R, with the package of ‘olsrr’ used to compare and select the best fitted model.^12^

### RNA-seq data and analysis

RNA-seq read counts and normalized expression (i.e., fragments per kilobase of nucleotides of transcripts per million of reads (FPKM)) data for stomach adenocarcinoma (STAD) patients and colon adenocarcinoma (COAD) patients were downloaded the website for The Cancer Genome Atlas (TCGA) via Genomic Data Commons (GDC) Data Portal (https://portal.gdc.cancer.gov/). Sample type information (normal vs. adenocarcinoma) was parsed from ‘file_id’ for each data file with ‘UUIDtoBarcode’ from the R package TCGAutils (version 1.8.0),^13^ with ‘01’ and ‘11’ in the submitter_id field of the code translated into ‘Normal’ and ‘STAD or ‘COAD’, respectively. Gene differential expression analysis was performed using methods within the generalized linear regression scheme implemented in the R package edgeR (version 3.30.3).^14^ Only STAD and COAD samples with matched normal tissues were used for this differential expression analysis. A STAD or COAD sample was matched with its normal sample collected from the same patients. Differential expression cutoff was set as absolute fold change >=2 and *q*-value<0.01, with *q*-value calculated using the R package qvalue (version 2.20.0).^15^ Gene set enrichment analysis was performed using the R package gage (version 2.37.0).^16^ List of genes participating innate immunity were downloaded from the database of InnateDB ^17^ (http://www.innatedb.com), and list of genes encoding cytokines were obtained from the article ^18^.

## Results

### Correlation of population age and sex ratio with COVID-19 mortality

As age and sex have been well-acknowledged as significant risk factors for poor outcomes with COVID-19,^3,4,7–9^ we first examined their correlations with COVID-19 mortality at the state level. We found no significant association of the proportion of old persons (age >65 years) with COVID-19 (Pearson’s *r*=-0.089, *p*=0.53; Table 1), suggesting ageism of a population does not necessarily imply increased vulnerability facing the pandemic. Surprisingly, we found a negative association between male-to-female population ratio and COVID-19 mortality (Pearson’s *r*=-0.47, *p*=5.5e-04). This was partially caused by higher inhabitation rate of women in metropolitan areas than men, as we observed a significant negative association between male-to-female ratio and population ratio of metropolitan areas (Pearson’s *r*=-0.45, *p*=9.8e-04), and a significant positive correlation for metropolitan population ratio with COVID-19 mortality (Pearson’s *r*=0.53, *p*=6.1e-05), at the state level. After correcting for metropolitan ratio with partial correlation calculation, the correlation between sex ratio and COVID-19 mortality was reduced though still significant (Partial Pearson correlation coefficient=-0.30, *p*=0.033).

**Table 1.**
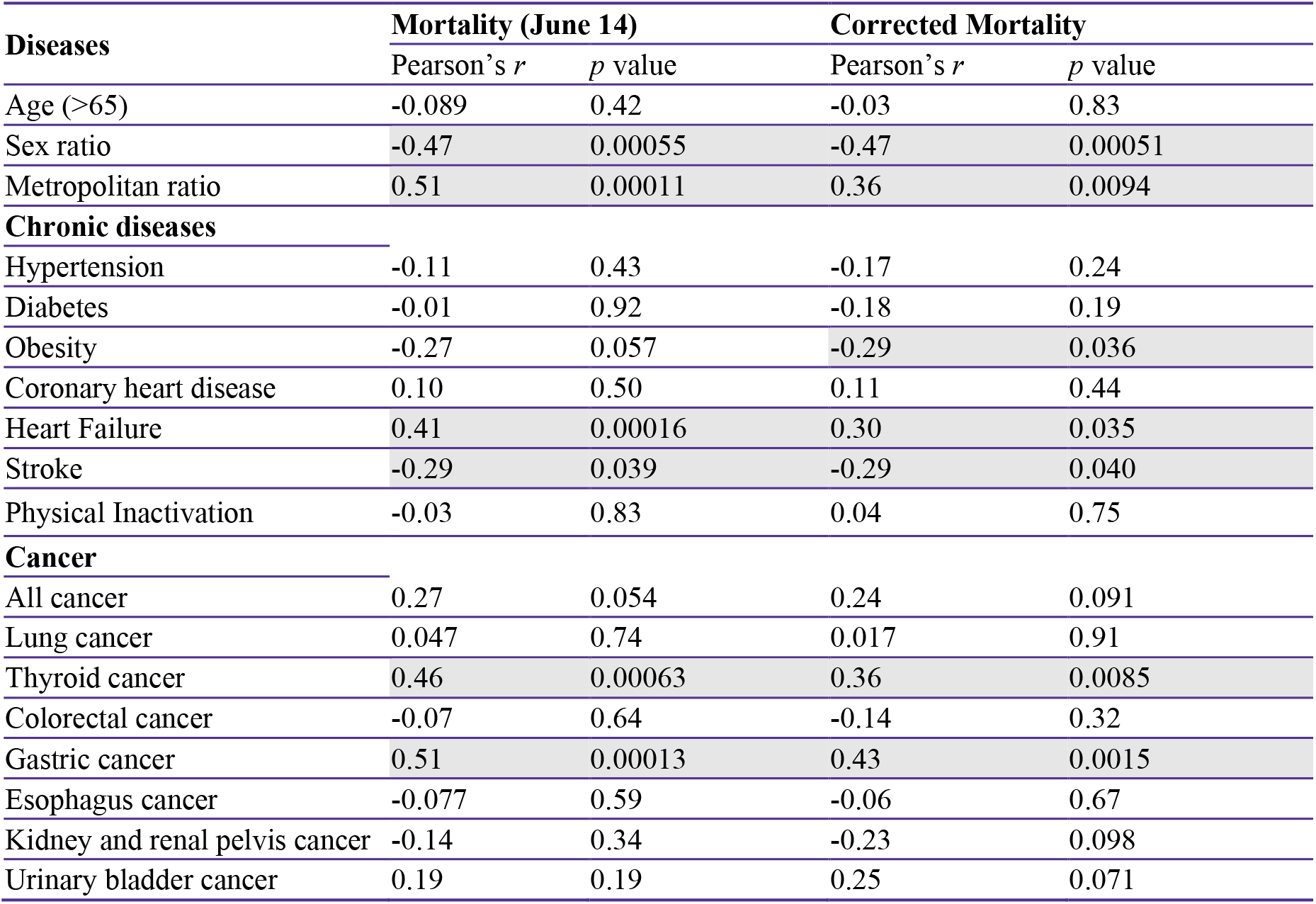
Correlation between prevalence of common chronic diseases and cancers with COVID-19 mortality for US states. Significant correlations (p<0.05) are highlighted in gray.

| Diseases | Mortality (June 14) |  | Corrected Mortality |  |
| --- | --- | --- | --- | --- |
| | Pearson's $r$ | $p$ value | Pearson's $r$ | $p$ value |
| Age (>65) | -0.089 | 0.42 | -0.03 | 0.83 |
| Sex ratio | -0.47 | 0.00055 | -0.47 | 0.00051 |
| Metropolitan ratio | 0.51 | 0.00011 | 0.36 | 0.0094 |
| **Chronic diseases** |  |  |  |  |
| Hypertension | -0.11 | 0.43 | -0.17 | 0.24 |
| Diabetes | -0.01 | 0.92 | -0.18 | 0.19 |
| Obesity | -0.27 | 0.057 | -0.29 | 0.036 |
| Coronary heart disease | 0.10 | 0.50 | 0.11 | 0.44 |
| Heart Failure | 0.41 | 0.00016 | 0.30 | 0.035 |
| Stroke | -0.29 | 0.039 | -0.29 | 0.040 |
| Physical Inactivation | -0.03 | 0.83 | 0.04 | 0.75 |
| **Cancer** |  |  |  |  |
| All cancer | 0.27 | 0.054 | 0.24 | 0.091 |
| Lung cancer | 0.047 | 0.74 | 0.017 | 0.91 |
| Thyroid cancer | 0.46 | 0.00063 | 0.36 | 0.0085 |
| Colorectal cancer | -0.07 | 0.64 | -0.14 | 0.32 |
| Gastric cancer | 0.51 | 0.00013 | 0.43 | 0.0015 |
| Esophagus cancer | -0.077 | 0.59 | -0.06 | 0.67 |
| Kidney and renal pelvis cancer | -0.14 | 0.34 | -0.23 | 0.098 |
| Urinary bladder cancer | 0.19 | 0.19 | 0.25 | 0.071 |

As the outbreak of the COVID-19 epidemic started asynchronously among different administrative regions, direct comparison for mortality between these regions could be biased. We thus calculated a corrected mortality (i.e., the COVID-19 mortality of a region on the 30^th^ day since the first day with at least 1 death in 100,000 people observed in that region). We obtained similar results with the corrected mortality (Table 1). The corrected mortality is used for subsequent analysis unless stated otherwise.

### Association of chronic disease prevalence with COVID-19 mortality

To find out if any population-level disease prevalence significantly correlates with the mortality of COVID-19, we calculated the Pearson’s correlation coefficients between each investigated chronic condition prevalence statistics and COVID-19 mortality at the US state level. Though hypertension, diabetes, and coronary heart disease were all reported as high incident comorbidities among COVID-19 patients with severe/death outcome by clinical cohort studies,^3,6–9,11,19–21^ we found no significant association between these factors and COVID-19 mortality at the state level (Table 1). Furthermore, the prevalence of stroke and physical inactivation were not related with COVID-19 mortality. Heart failure was found to be positively significantly correlated with COVID-19 mortality (Pearson’s *r*=0.36, *p*=0.013), and surprisingly, obesity was significantly negatively correlated with COVID-19 mortality (Pearson’s *r* =-0.42, *p*=0.0034; Table 1).

To investigate the potential confounding effects of these factors, we then performed a stepwise model selection analysis, with heart failure, obesity, stroke, coronary heart disease, diabetes, hypertension, age (>65), and sex ratio as the explanatory factors. The model with the lowest Mallow’s *Cp* was chosen which included heart failure, stroke, obesity, and sex ratio as independent predictors. Together, the 4 factors accounted for ∼51% of the variation in COVID-19 mortality among US states (*p*=6.7e-09). In this concise model, heart failure (HF) remained to be significant (*p*=0.0016), while obesity was not statistically insignificant (*p*=0.072). Notably, stroke became significant (*p*=0.0098) after controlling for the effects of obesity and perhaps heart failure. This result suggests that the significant negative correlation for obesity prevalence with COVID-19 mortality was more likely caused by the negative association of stroke with COVID-19, since obesity is a known risk factor for stroke,^22^ and significantly positively correlated with each other (Pearson’s *r*=0.67, *p*=9.1e-08).

### Cancers associated with COVID-19 mortality

We then analyzed the contributions of cancer rate to the regional variations in COVID-19 mortality. No significant association was found for the incidence rate of all cancers with COVID-19 mortality (Pearson’s *r*=0.26; *p*=0.082), nor for the incidence of lung cancer (Pearson’s *r*=-0.011; *p*=0.94). However, we found significant correlation with COVID-19 mortality for the incidence rate of thyroid cancer (Pearson’s *r*=0.49; *p*=4.e3-04) and gastric cancer (Pearson’s *r*=0.61; *p*=5.2e-06).

To disentangle potential confounding effects among different cancers, we performed a stepwise model selection analysis to test all the possible combinations of the investigated cancers. The model with the lowest Mallow’s *Cp* was selected, which included gastric cancer (*p*=8.7e-06), thyroid cancer (*p*=1.9e-03), and other cancers (*p*=0.43) as independent predictors for the state-level COVID-19 mortality (Table 3). These factors together accounted for ∼50% of the variation observed in COVID-19 mortality across US states (*p*=3.5e-07).

**Table 2.** Summary of the multivariate linear model for chronic diseases.

| **Diseases** | **Mortality (June 14)** |  | **Corrected Mortality** |  |
| --- | --- | --- | --- | --- |
|  | Coefficient | Pr(>\|t\|) | Coefficient | Pr(>\|t\|) |
| Sex | -2.76 | 0.081 | -1.21 | 0.11 |
| Heart Failure | 5.40 | 0.00016 | 1.81 | 0.0016 |
| Stroke | -20.15 | 0.0017 | -7.01 | 0.0098 |
| Obesity | -2.75 | 0.024 | -0.94 | 0.072 |

**Table 3.** Summary of the multivariate linear model for cancers.

| **Diseases** | **Mortality (June 14)** |  | **Corrected Mortality** |  |
| --- | --- | --- | --- | --- |
|  | Coefficient | Pr(>\|t\|) | Coefficient | Pr(>\|t\|) |
| Gastric cancer | 13.90 | 1.1e-04 | 6.29 | 8.7e-06 |
| Thyroid cancer | 5.68 | 2.2e-03 | 2.01 | 1.9e-03 |
| Other cancers | 0.15 | 0.22 | 0.04 | 0.43 |

### RNA-seq analysis about stomach cancer

Carcinogenesis is a complex biological process depending on numerous genetic and environmental factors.^23^ As both gastric cancer (GC) and thyroid cancer (TC) are not highly prevalent in the United States (Incidence rate for GC: 6/100,000; that for TC: 13/10000), the correlations between the incidence rate of gastric cancer and thyroid cancer with COVID-19 mortality were likely due to underlying factors, such as environment and lifestyle, especially risk factors leading to these two cancers. Moreover, except for a few cases, including lung cancer, female breast cancer, cervical cancer, colon and rectum cancer, there are currently no effective screening method and diagnostic technique for most early-state cancers or precancerous diseases.^24^ It is thus likely that a lot more people might be exposed to risk factors for a cancer than indicated by the incidence rate. Risk factors imposed by certain environmental conditions and lifestyles were transformed into endogenous genetic responses which first induced chronic inflammation, such as gastritis, and finally developed into cancer. As a final state of these transformations, the carcinogenic transcriptome not only summarizes the accumulated genetic responses, but also indicates clues for the mechanisms of carcinogenesis from which targeted preventive measures could be developed. We thus analyzed the transcriptome of stomach adenocarcinoma (STAD), which is the most common type of stomach cancer,^25^ against the transcriptome of colon carcinoma (COAD).

We downloaded the raw read counts normalized gene expression data of the RNA-seq experiments for STAD and COAD from The Carcinoma Genome Atlas (TCGA) project. We first compared the expression levels for Angiotensin-converting Enzyme 2 (ACE2) and Transmembrane protease, serine 2 (TMPRSS2), which have been identified as the primary receptor and the priming factor for the entry of SARS-Cov-2 into host cells, respectively.^26(p2)^ The expression levels of ACE2 and TMPRSS2 were down-regulated in the two carcinomas compared to the healthy tissues (Figure 2).

**Figure 2.**
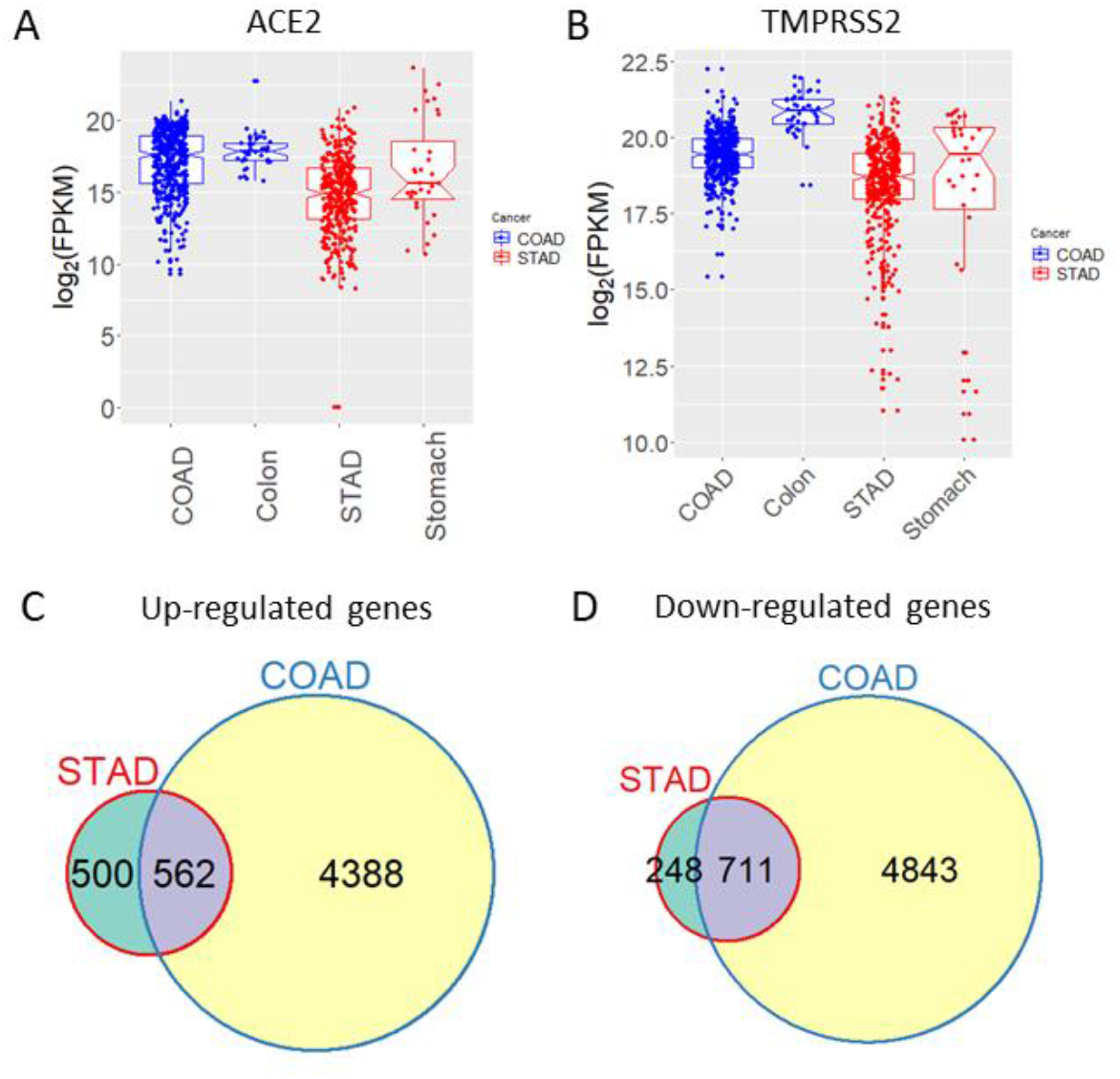
Comparative expression analysis for STAD and COAD. Normalized expression in STAD, COAD, healthy stomach, and healthy colon for ACE2 (A) and TMPRSS2 (B). Comparison between up-regulated (C) down-regulated (D) genes in STAD and COAD carcinomas.

We then performed gene differential analysis separately for 27 pairs of matched STAD and normal stomach tissues, and 41 pairs of matched COAD and normal colon tissues. We identified 1,062 and 4,950 up-regulated genes and 959 and 5,554 down-regulated genes for STAD and COAD, respectively (Figure 2). The comparison showed, for STAD, ∼53% and ∼74% of the up-regulated and down-regulated genes, respectively, were also correspondingly regulated in COAD. Interestingly, both ACE2 and TMPRSS2 were significantly down-regulated in STAD, with the expression of ACE2 reduced to 8.6% of that in the normal stomach (*q*-value=1.89e-33) and TMPRSS2 lowered to 64% of the normal levels (*q*-value=3.83e-03). Both genes were also significantly repressed in COAD, however, with TMPRSS2 down-regulated to a much lower level than ACE2 (ACE2: fold change=0.71, *q*-value=0.019; TMPRSS2: fold change=0.39, *q*-value=3.76e-37).

Despite the much larger transcriptomic alterations in COAD versus STAD, gene set enrichment analysis (GSEA) using Gene Ontology (GO) annotation data identified more interesting scenarios for STAD than for COAD. We found that, immune responses related pathways were broadly more highly up-regulated or less down-regulated in STAD than in COAD, implicating a systematically more active immune activities in STAD compared to COAD (Table 4). In comparison, basic metabolism pathways were less up-regulated or more down-regulated in STAD than in COAD (Table 5). We obtained similar results by GSEA using the annotation data for human genes in the database of Kyoto Encyclopedia of Genes and Genomes (Table 6 and Table 7). Examination of the expression changes of genes participating in immune responses found that, genes involved in innate immunity were more dramatically up- or down-regulated in STAD than in COAD (Figure 3A and 3B), whereas genes encoding cytokines were up-regulated in STAD than in COAD to larger extent, but were down-regulated in both adenocarcinoma to the same extent (Figure 3C and 3D). These findings suggested immune responses were more significantly altered in STAD than in COAD, implying higher severity regarding to inflammation and/or malignancy, consistent with much poorer survival rates of stomach cancer relative to colon cancer (All stages combined 5-year survival rates for stomach cancer: 32%, that for colon cancer: 67%; data source: https://www.cancer.org/, website of the American Cancer Society).

**Table 4.** Biological processes with more positive regulation in STAD than in COAD.

| GO ID | Dif. regulation | *q* value | Gene number | Term |
| --- | --- | --- | --- | --- |
| GO:0002682 | 6.08 | 3.8E-07 | 269 | regulation of immune system process |
| GO:0002684 | 5.77 | 1.6E-06 | 151 | positive regulation of immune system process |
| GO:0001775 | 5.19 | 1.6E-05 | 270 | cell activation |
| GO:0002250 | 4.88 | 7.7E-05 | 95 | adaptive immune response |
| GO:0002460 | 4.87 | 7.7E-05 | 94 | adaptive immune response based on somatic recombination of immune receptors built from immunoglobulin superfamily domains |
| GO:0002252 | 4.63 | 1.5E-04 | 146 | immune effector process |
| GO:0002449 | 4.55 | 2.3E-04 | 87 | lymphocyte mediated immunity |
| GO:0002694 | 4.32 | 4.7E-04 | 114 | regulation of leukocyte activation |
| GO:0002443 | 4.31 | 4.7E-04 | 98 | leukocyte mediated immunity |
| GO:0002521 | 4.06 | 1.0E-03 | 151 | leukocyte differentiation |
| GO:0002520 | 4.02 | 1.0E-03 | 254 | immune system development |
| GO:0002504 | 4.34 | 3.4E-03 | 16 | antigen processing and presentation of peptide or polysaccharide antigen via MHC class II |
| GO:0002253 | 3.71 | 3.9E-03 | 65 | activation of immune response |
| GO:0002696 | 3.60 | 5.1E-03 | 71 | positive regulation of leukocyte activation |
| GO:0005975 | 3.31 | 1.0E-02 | 478 | carbohydrate metabolic process |
| GO:0001816 | 3.27 | 1.2E-02 | 147 | cytokine production |
| GO:0002541 | 3.08 | 2.6E-02 | 39 | activation of plasma proteins during acute inflammatory response |
| GO:0001906 | 3.14 | 2.6E-02 | 25 | cell killing |
| GO:0002697 | 3.00 | 2.7E-02 | 65 | regulation of immune effector process |
| GO:0002455 | 3.03 | 2.9E-02 | 33 | humoral immune response mediated by circulating immunoglobulin |

**Table 5.** Biological processes with more negative regulation in STAD than in COAD.

| **GO ID** | **Dif. regulation** | **q value** | **Gene Number** | **Term** |
| --- | --- | --- | --- | --- |
| GO:0006414 | -7.11 | 1.1E-08 | 99 | translational elongation |
| GO:0006412 | -5.59 | 2.8E-06 | 367 | translation |
| GO:0006396 | -3.66 | 1.4E-02 | 499 | RNA processing |
| GO:0006364 | -3.57 | 2.2E-02 | 81 | rRNA processing |

**Table 6.**
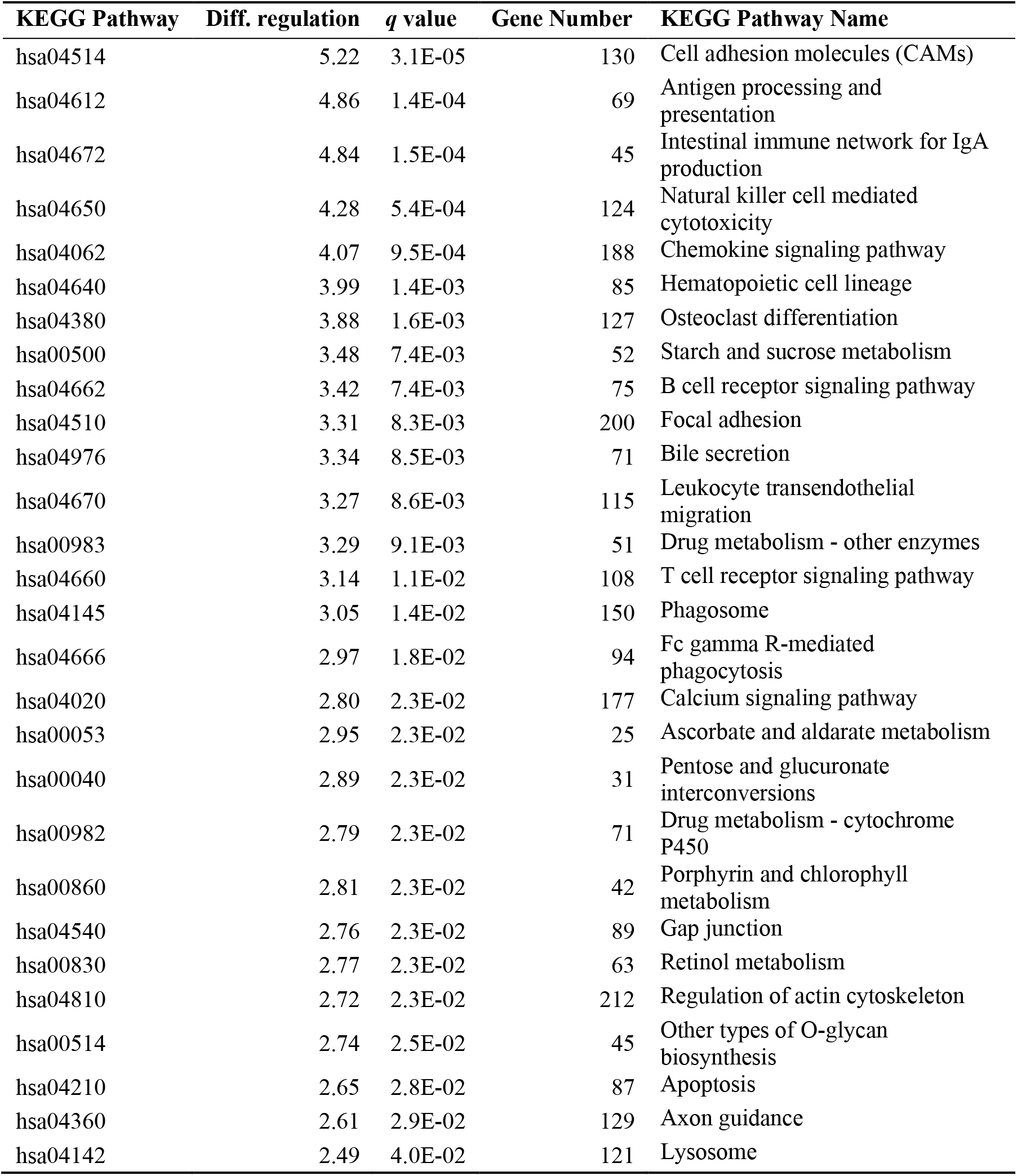
KEGG pathways with more positive regulation in STAD than in COAD.

| KEGG Pathway | Diff. regulation | *q* value | Gene Number | KEGG Pathway Name |
| --- | --- | --- | --- | --- |
| hsa04514 | 5.22 | 3.1E-05 | 130 | Cell adhesion molecules (CAMs) |
| hsa04612 | 4.86 | 1.4E-04 | 69 | Antigen processing and presentation |
| hsa04672 | 4.84 | 1.5E-04 | 45 | Intestinal immune network for IgA production |
| hsa04650 | 4.28 | 5.4E-04 | 124 | Natural killer cell mediated cytotoxicity |
| hsa04062 | 4.07 | 9.5E-04 | 188 | Chemokine signaling pathway |
| hsa04640 | 3.99 | 1.4E-03 | 85 | Hematopoietic cell lineage |
| hsa04380 | 3.88 | 1.6E-03 | 127 | Osteoclast differentiation |
| hsa00500 | 3.48 | 7.4E-03 | 52 | Starch and sucrose metabolism |
| hsa04662 | 3.42 | 7.4E-03 | 75 | B cell receptor signaling pathway |
| hsa04510 | 3.31 | 8.3E-03 | 200 | Focal adhesion |
| hsa04976 | 3.34 | 8.5E-03 | 71 | Bile secretion |
| hsa04670 | 3.27 | 8.6E-03 | 115 | Leukocyte transendothelial migration |
| hsa00983 | 3.29 | 9.1E-03 | 51 | Drug metabolism - other enzymes |
| hsa04660 | 3.14 | 1.1E-02 | 108 | T cell receptor signaling pathway |
| hsa04145 | 3.05 | 1.4E-02 | 150 | Phagosome |
| hsa04666 | 2.97 | 1.8E-02 | 94 | Fc gamma R-mediated phagocytosis |
| hsa04020 | 2.80 | 2.3E-02 | 177 | Calcium signaling pathway |
| hsa00053 | 2.95 | 2.3E-02 | 25 | Ascorbate and aldarate metabolism |
| hsa00040 | 2.89 | 2.3E-02 | 31 | Pentose and glucuronate interconversions |
| hsa00982 | 2.79 | 2.3E-02 | 71 | Drug metabolism - cytochrome P450 |
| hsa00860 | 2.81 | 2.3E-02 | 42 | Porphyryn and chlorophyll metabolism |
| hsa04540 | 2.76 | 2.3E-02 | 89 | Gap junction |
| hsa00830 | 2.77 | 2.3E-02 | 63 | Retinol metabolism |
| hsa04810 | 2.72 | 2.3E-02 | 212 | Regulation of actin cytoskeleton |
| hsa00514 | 2.74 | 2.5E-02 | 45 | Other types of O-glycan biosynthesis |
| hsa04210 | 2.65 | 2.8E-02 | 87 | Apoptosis |
| hsa04360 | 2.61 | 2.9E-02 | 129 | Axon guidance |
| hsa04142 | 2.49 | 4.0E-02 | 121 | Lysosome |

**Table 7.** KEGG pathways with more negative regulation in STAD than in COAD.

| KEGG Pathway | Diff. Regulation | *q* value | Gene Number | KEGG Name |
| --- | --- | --- | --- | --- |
| hsa03010 | -7.18 | 6.1E-09 | 88 | Ribosome |
| hsa03013 | -2.79 | 2.4E-01 | 151 | RNA transport |
| hsa03008 | -2.26 | 7.1E-01 | 74 | Ribosome biogenesis in eukaryotes |
| hsa00100 | -2.11 | 7.8E-01 | 19 | Steroid biosynthesis |
| hsa03040 | -1.97 | 7.8E-01 | 127 | Spliceosome |
| hsa03020 | -1.97 | 7.8E-01 | 29 | RNA polymerase |
| hsa03018 | -1.79 | 9.0E-01 | 71 | RNA degradation |

**Figure 3.**
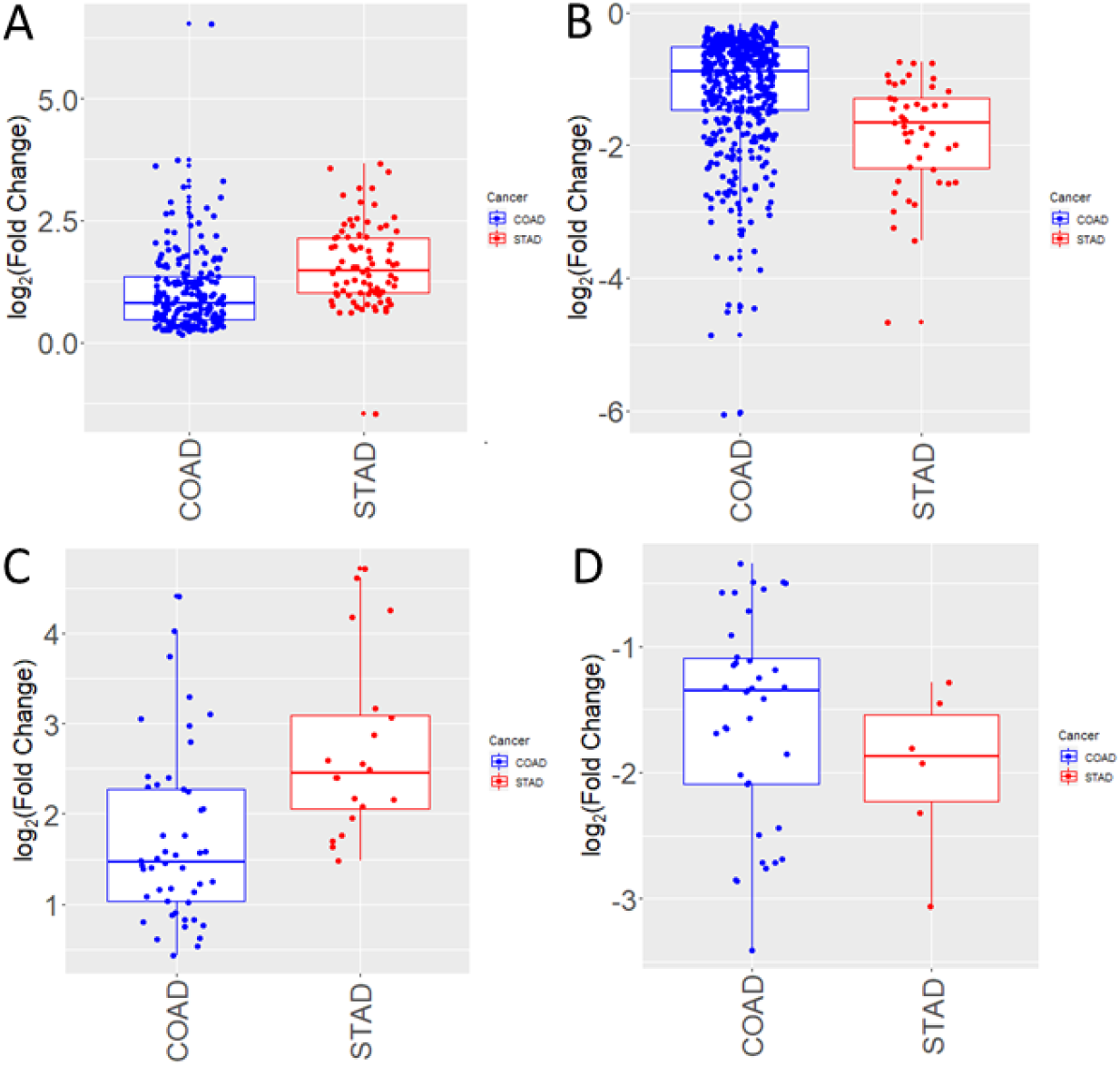
Comparison of differentially expressed innate immune response and cytokine genes between STAD and COAD. A) Innate immune genes up-regulated in STAD and COAD; B) Innate immune genes down-regulated in STAD and COAD; C) Cytokine genes up-regulated in STAD and COAD; D) Cytokine genes down-regulated in STAD and COAD.

## Discussion

In this study, we conducted a population-level association analysis between the prevalence/incidence rate of a group of common chronic diseases and cancers with COVID-19 mortality, for US states and counties. Our results implicated specific concerns towards particular diseases, in regarding to the management of public health crisis caused by an epidemic. Specifically, it was shown that, the rate of heart failure, stomach cancer, and thyroid cancer could explain a substantial proportion of the regional variations observed for COVID-19 mortality.

Heart failure has been discovered as the leading risk factor among comorbidities for adverse outcome of COVID-19.^4,7,11,27^ Interestingly, obesity was negatively correlated with COVID-19 mortality but was insignificant in the multivariate model with stroke added. The result suggest at the population level the influence of obesity on COVID-19 outcomes may be through that of stroke, as obesity is one of the major risk factor for stroke.^22^ The negative correlation for stroke/obesity could be explained by the common prescription of anti-coagulant drugs to patients with obesity or stroke, since blood coagulation and thrombosis have been found to be related with poor prognosis of COVID-19 patients.^28–32^ The population-level positive association between heart failure with COVID-19 mortality might represent combined effects of heart failure and of the risk factors for heart failure, including age, sex, coronary artery disease, myocardial infarction, hypertension, diabetes, and obesity.^33–35^ Therefore, once infected with COVID-19, patients with these risk factors for heart failure require closer monitoring to prevent events of heart failure.

We also identified thyroid cancer and gastric cancer as potential predictive factors for COVID-19 mortality. The associations between the prevalence of specific types of cancers with COVID-19 mortality could be indirect, more likely a manifestation of underlying risk factors, such as environment conditions and lifestyle. Acknowledged risk factors for gastric cancer including poor diet, lifestyle (e.g., alcohol consumption, smoking, inactivity), genetics, *Helicobacter pylori* infection, age (>50 years), and sex (male).^25,36,37^ *H. pylori* infection is considered the major risk factor for non-cardia gastric carcinoma (NCGA).^25,38,39^ Approximately 50% of the world’s population are infected by *H. pylori*.^40^ Colonization of *H. pylori* in the gastric mucosa causes a systemic immune response, chronically forming an increasingly inflamed environment, favoring development of corpus-predominant gastritis, gastric atrophy, and intestinal metaplasia.^40,41^ In addition, the systematically activated immune response results in increased production of reactive oxygen and nitrogen species which can subsequently cause cell and DNA damage, inducing oncogenic mutations facilitating carcinogenesis.^41^ While previously thought to be unrelated or reverse related, *H. pylori* can also play as risk factors for cardia gastric carcinoma (CGA) in high-risk situations.^38^ Specific risk factors for CGA include gastroesophageal reflux disease and hiatal hernia, which in turn suffer risks from similar demographic and lifestyle factors as NCGA and CGA.^25^ Previous transcriptome studies have identified increased activities in genes encoding for pro-inflammatory and anti-inflammatory factors, including cytokines and chemokines, in gastritis and early and late stage gastric cancers.^42–44^ Consistently our analysis showed that immune responses in stomach adenocarcinoma, which is the dominant type of gastric cancer, were more active than in colon adenocarcinoma, possibly because the environment in the colon is less inflammatory. The much stronger repression of ACE2 in STAD than in COAD might be one of the responsible factor for the much poorer prognosis of STAD than COAD, since ACE2 has been found to have anti-tumor roles,^45^ and high expression of ACE2 in patients with endometrial carcinoma and renal papillary carcinoma is linked with good prognosis. ^46^ In relation to COVID-19 transmission, this fact indicates higher risk of STAD patients than COAD patients, since ACE2 acts in protective roles for multiple organs along with being the primary receptor for the entry of SARS-Cov-2 into host cells.^47^ Taken together, these facts implicate that COVID-19 patients with comorbidities that are characterized by more severely downgraded inflammatory status or remarkably down-regulated ACE2 expression could have higher risks for adverse outcomes.

The last decades have seen a worldwide continuous increase in the incidence of thyroid cancer.^48–50^ While this could partially be ascribed to improved detection rate,^51^ actual increases cannot be ruled out and changes in risk factors have been acknowledged.^50,52^ Leading risk factors in consideration include increased radiation exposure due to over-diagnosis by radiation-imposing methods (e.g., X-ray, CT scan), universal salt iodization, lifestyle factors (dietary nitrate, smoking), obesity, and estrogen.^52^ However, more studies are needed to unravel the mechanisms linking these factors with increased incidence of thyroid cancer.

The major limitation of our study is the limited public health, demographic, as well as epidemic data, in both depth and breadth. More systematic analysis using public health and epidemiological data from more countries and regions, and prevalence/incidence data for more diseases, is needed to validate and expand our findings. Despite intensive studies and numerous reports about the risks, mechanisms, intervention adaptation for comorbidities (especially heart diseases) in terms of clinical strategy for constraining damages caused by COVID-19, relatively fewer attention and efforts have been paid on the risks of patients with cancers and subclinical conditions related the development of cancers. Our study suggests more attention to be paid on excess COVID-19 mortality caused by cancer-related health conditions.

## Conclusion

Population data on chronic health conditions were used to examine the potential links between comorbidities to COVID-19 mortality. The results presented herein support an increased risk of mortality with heart failure, thyroid cancer, and gastric cancer. While we have evaluated potential pathways and causes to explain differences in fatality, further studies including well-conceived clinical trials need to be conducted to both verify a relationship in these comorbidities and examine the pathways of action.

## Data Availability

All data are available upon request.

